# Cost Comparison and Spending on Tobacco Products: Evidence from A Nationally Representative Sample of Adult E-Cigarette Users

**DOI:** 10.1101/2024.04.03.24305296

**Authors:** Shaoying Ma, Qian Yang, Sooa Ahn, Hojin Park, Yanyun He, John F P Bridges, Ce Shang

## Abstract

**Background:** Over 20 states and local jurisdictions in the U.S. have imposed e-cigarette taxes. It is important to evaluate how adult vapers, including those who also smoke respond to e-cigarette taxation. The purpose of this study is to examine factors associated with adult vapers’ cost perception of e-cigarettes relative to cigarettes and budget allocations between two products.

**Methods:** We recruited a nationally representative sample of 801 adult e-cigarette users in the U.S., who participated in an online survey in April and May 2023. Nested-ordered logit models and ordinary least squares regressions were used in the analysis.

**Results:** On average, monthly e-cigarette spending was $82.22, and cigarette spending was $118.77 among dual users. Less frequent e-cigarette use and higher state-level e-cigarette taxes were associated with perceiving smoking as cheaper than vaping. Age and exclusive use of tank systems were associated with perceiving vaping as cheaper than smoking. Exclusive use of tank systems was also associated with lower e-cigarette spending. Adults who used e-cigarettes more frequently preferred to report weekly budget on e-cigarettes (*p* < 0.01), and among dual users, everyday smokers preferred to report weekly (versus monthly) budget on cigarettes compared to someday smokers (*p* < 0.001).

**Conclusion:** Among US adult vapers, frequencies of tobacco use and e-cigarette device type are closely related to cost measures; and e-cigarette taxes are associated with cost perception of e-cigarettes relative to cigarettes, suggesting potential financial disincentive for vaping. Policymaker may consider imposing differential taxes by e-cigarette product types due to their different costs to consumers.

## INTRODUCTION

In 2021, 4.5% US adults aged 18 or above were current e-cigarette users, including 11% of young adults aged 18-24.(Kramarow & Elgaddal, 2023) While e-cigarettes lead to addiction among youth and young adults, they are also considered as a less harmful alternative to combustible tobacco products, thereby having the potential to help quitting smoking if smokers switch completely to e-cigarettes.(Balfour et al., 2021; National Academies of Sciences Engineering and Medicine, 2018) As the popularity of e-cigarettes grows over time,(Centers for Disease Control and Prevention (CDC), 2023b) it becomes increasingly important to understand adults e-cigarette users’ purchase and use behaviors in order to inform policymakers who are balancing the benefits and risks of e-cigarettes.

In particular, cost comparison between e-cigarettes and cigarettes and the spending on each product are particularly relevant to the understanding of adult e-cigarette purchase and use behaviors. Approximately 30% of adult e-cigarette users in the U.S. are dual users who also concurrently smoke cigarettes and 40% are former smokers who have smoked 100 cigarettes or more in the past.(Centers for Disease Control and Prevention (CDC), 2023a). While 30% of adult e-cigarettes never smoked, they are more likely to be young adults - a group that may be exposed to the gateway effect where e-cigarette use leads to future smoking.(He et al., 2024; Takada et al., 2022; Yao et al., 2017) Therefore, the budget or expenditures on tobacco products and the cost comparison reveal economic incentives and tradeoffs when choosing between two products, further influencing downstream behavioral outcomes including quitting and transitions (e.g., escalation, relapse).

How e-cigarette users consider cost comparison between cigarettes and e-cigarette and their spending on tobacco products may change as a growing number of states and local jurisdictions tax e-cigarettes. Using a nationally representative sample of smokers and vapers from the U.S., one study found that higher frequency of cigarette use leads smokers and vapers to think cigarettes are more expensive and e-cigarette users who use pre-filled cartridges and tank devices are more likely to think e-cigarettes are less expensive than cigarettes.(Thompson et al., 2019) However, given the small sample size of vapers in that study, their conclusions may be largely driven by smokers, with a limited ability to inform e-cigarette users’ perceived relative cost.

In addition to cost consideration, income-related factors such as expenditures and budgets are also important factors of consumers decision-making. One study from the U.S. suggests that expenditures could be a useful measure of e-cigarette use intensities among smokers, which are often not captured in detail in national surveys. Studies from the UK further illustrate that expenditures on e-cigarettes change as the marketplace evolves and are associated with product choices, use behaviors, and demographics. However, evidence on tobacco and e-cigarette expenditures for the US population is scarce.

Motivated by these knowledge gaps, our study contributes to the tobacco control literature by evaluating the expenditures on cigarettes and e-cigarettes (frequencies and amounts), and the cost comparison between two products, among a nationally representative sample of adult e-cigarette users aged 18+. We further assessed how expenditures and cost comparisons are associated with taxation, use behaviors, and socio-demographic characteristics.

## METHOD

### Eligibility and sample size

We recruited a nationally presentative sample of 808 adult vapers in the U.S. through Ipsos KnowledgePanel in April and May 2023. The eligibility criteria were: 1) at least 18 years old; 2) have used e-cigarettes in the past 30 days; and 3) frequency of e-cigarette use is either every day or some days. The sample was weighted to represent e-cigarette users aged 18+ using the 2021 National Health Interview Survey (NHIS) as benchmark and based on the following characteristics: sex by age, race/ethnicity, census region, metropolitan status, education, and vaping and smoking frequencies. In the analysis, we excluded 4 participants who did not report e-cigarette device types and 3 participants who refused to answer the cost comparison question. Therefore, the survey data from a total of 801 participants were utilized in this study.

Given that we estimated factors associated with spending and cost comparison, participants who reported zero spending on either product (cigarette or e-cigarette) were excluded from the analysis, except for exclusive e-cigarette users, whose total expenditures equaled their e-cigarette expenditures. A total of 776 participants were included in the analysis of e-cigarette expenditures and total expenditures, whereas 268 dual users of cigarettes and e-cigarettes were included in the cigarette expenditure analysis.

## Measures

### Outcome measures (dependent variables)

#### Cost comparison

Study participants were asked how they compared the cost of vaping to smoking, i.e., e-cigarettes are less expensive than cigarettes, about the same, or more expensive, or they simply did not know. Conditional on participants not choosing “don’t know”, we constructed an order variable where higher value represented e-cigarettes to be more expensive than cigarettes (1= “e-cigarettes are less expensive”, 2 = “two are about the same”, and 3 = “e-cigarettes are more expensive”).

#### Expenditure

Participants reported their spending on e-cigarettes in dollars on a weekly or monthly basis to their liking, and if they were current smokers, their spending on cigarettes. Specifically, adult e-cigarette users were asked the following: “We’d like to find out how much you spend on e-cigarettes regularly. Is it easier for you to say how much you spend on e-cigarettes per week or how much you spend on e-cigarettes per month?” Dual users of e-cigarettes and cigarettes were asked a similar question on cigarette spending. Using this information, we constructed indicators for reporting weekly versus monthly expenditures on vaping and smoking, respectively. To convert weekly expenditures into standardized monthly expenditures, we multiplied weekly expenditures by 4. We also calculated each dual user’s total monthly expenditures on e-cigarettes and cigarettes. These monthly expenditure measures (e-cigarette, cigarette, and two products combined) were transformed into the logarithmic form.

### Explanatory variables

#### Tobacco use

Tobacco use patterns in the past 30 days were asked in the survey and measured by the frequency of vaping e-cigarettes (some days or every day) and the frequency of smoking cigarettes (not at all, some days, or every day), which were used as explanatory variables in our analysis.

#### Product type

We also controlled for e-cigarette product type used in the past 30 days based on survey questions: exclusively disposables, exclusively rechargeable tank systems with e-liquids, exclusively rechargeable systems with pre-filled pods or cartridges, and multiple types of e-cigarette products.

#### Tobacco taxation

State-level cigarette and e-cigarette taxes as of December 2022 were obtained from the CDC’s State Tobacco Activities Tracking and Evaluation (STATE) System. Cigarette taxes were measured in the unit of US dollars per pack. Following existing literature, we used a standardization method to convert ad valorem e-cigarette taxes into specific tax per 1 ml of e-liquid volume, assuming a retailer to wholesaler markup rate of 35%.(Cotti et al., 2021)

#### Socio-demographics

We controlled for socio-demographic variables in all specifications, including participant age (30-44, 45-59, 60+, 18-29 as reference), sex (female, male as reference), race/ethnicity (Non-Hispanic Black, Non-Hispanic other race, Hispanic, non-Hispanic White as reference), marital status (married, not married as reference), number of adults in the household, housing type (condo/townhouse attached, apartment complex, other, one-family house as reference), education attainment (high school, some college or associate degree, bachelor’s degree or more, less than high school as reference), household income ($25K to less than $50K, $50K to less than $75K, $75K or more, less than $25K as reference), and employment status (working part-time, not working, working full-time as reference).

### Analysis

#### Cost comparison analysis

##### Cost comparison analysis

Since cost comparison question allowed for “don’t know” option, our analysis used a two parts model: first, we estimated a logit model (Equation 1) to obtain the predicted probability that a person knew the relative price, i.e., they did not choose “don’t know”; second, we estimated an ordered logit model (Equation 2) by regressing cost comparison on explanatory variables while controlling for the predicted probability of knowing the relative price. This two-part model performed better than alternative multinomial regressions according to Akaike information criterion (AIC) and Bayesian information criterion (BIC).

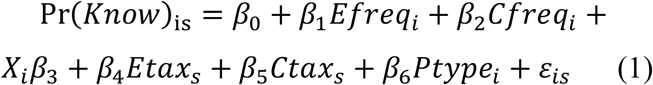

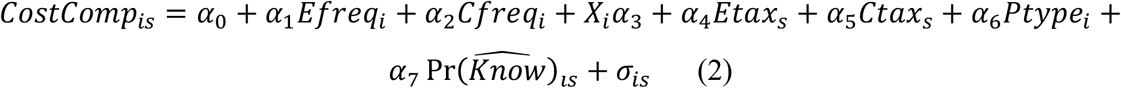

In Equation 1, Pr(Know)_*is*_ equals 1 if participant *i* from state *s* answered, “e-cigarette are less expensive”, “about the same”, or they are “more expensive” than cigarettes, and Pr(Know)_*is*_ equals 0 if participant *i* answered “don’t know”. *Efreq*_*i*_ and *Cfreq*_*i*_ are frequencies of e-cigarette and cigarette use, respectively. *X*_*i*_ is a vector of socio-demographic variables. *Etax*_*s*_ is standardized e-cigarette tax per ml in US dollars (see “Tobacco taxation” subsection above for more details), and *Ctax*_*s*_ is defined as cigarette tax per pack in US dollars. *Ptype*_*i*_ is the e-cigarette product type that participant used in the past 30 days.

Since *CostComp*_*is*_ is an ordinal outcome measure, the model was estimated using ordered logit. We obtained the odds ratio for each independent variable, which represented the change in the odds of *CostComp*_*is*_ associated with a one-unit change in the corresponding independent variable, holding the other independent variables constant. 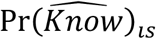 is the predicted probability we obtained from step one.

##### Expenditure analysis

To understand participants’ cost comparison, we first compared their preference for reporting weekly vs. monthly expenditures by use status (dual user vs. exclusive e-cigarette user, and every day vs. someday e-cigarette use). Next, in Equation 3, we examined the associations between tobacco expenditures and frequencies of use, product type, socio-demographics, and tobacco taxation. Equation 3 was estimated for three different expenditure measures: expenditures on e-cigarettes, expenditures on cigarettes, and the sum of those two (i.e., total expenditure). In Equation 4, we further included cost comparison in the analysis to assess whether cost comparison mediated the associations between taxes and expenditures. For all analyses, standard errors were clustered at the state level.

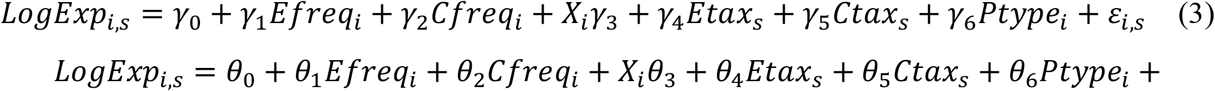

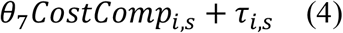

## RESULTS

Table 1 presents the summary statistics of outcome variables, tobacco use frequencies, product type, and state-level taxes. Among adult e-cigarette users in the U.S., 50% were everyday users and 50% were someday users; more than two thirds (68.92%) were exclusive e-cigarette users who never smoked or were not current smokers. 42% perceived e-cigarettes as less expensive than cigarettes, 24% reported perceiving the costs of cigarettes and e-cigarettes as about the same, 18% believed e-cigarettes were more expensive, and 17% said that they did not know. Average monthly e-cigarette spending was $82.22 and average monthly cigarette spending was $118.77. Standardized state e-cigarette taxes per ml were 0.54 dollars on average and state cigarette tax was 1.85 dollars per pack at the time of survey.

**Table 1.** Summary statistics of dependent and independent variables (socio-demographic variables in Appendix)

| *Variable* | Frequency | Weighted<br>percentage/mean |
| --- | --- | --- |
| *Cost comparison* |  |  |
| E-cigarettes are less expensive than cigarettes | 418 | 41.86% |
| About the same | 187 | 23.69% |
| E-cigarettes are more expensive than cigarettes | 115 | 17.81% |
| Don' know | 81 | 16.65% |
| *Converted monthly expenditure (dollars)* |  |  |
| E-cigarettes | 776 | 82.22 |
| Cigarettes | 268 | 118.77 |
| *E-cigarette frequency of use* |  |  |
| Some days | 241 | 49.91% |
| Everyday | 560 | 50.09% |
| *Cigarette frequency of use* |  |  |
| Not at all | 520 | 68.92% |
| Some days | 118 | 15.13% |
| Everyday | 163 | 15.95% |
| *Product type of use* |  |  |
| Disposable only | 219 | 33.22% |
| E-liquids only | 261 | 27.70% |
| Pre-filled pods only | 141 | 13.28% |
| Multiple | 180 | 25.80% |
| State e-cigarette tax (dollar per ml) | 801 | 0.54 |
| State cigarette tax (dollar per pack) | 801 | 1.85 |
| *N* | 801 |  |

Frequency column is the number of observations in each category and weighted percentage/mean column presents weighted percentage of categorical variables and weighted mean of continuous variables.

Appendix Table 1 presents the summary statistics of demographic characteristics. The majority of e-cigarette users were young people aged 18-29 (43%) and 30-44 (33%), male (54%), unmarried (68%), non-Hispanic White (73%), working full-time (54%), had a household income of $75K or less (56%), lived in a one-family house (65%), and had some college education or associate degree or less (79%).

Table 2 presents our findings from the cost comparison analysis. Relative to someday e-cigarette users, everyday e-cigarette users were less likely to perceive smoking as cheaper (OR = 0.55, *p* < 0.05). Compared to young adults aged 18-29, those who were at least 45 years old were less likely to perceive smoking as cheaper (OR = 0.23, *p* < 0.01 for 45-59 and OR = 0.32, *p* < 0.05 for 60+). Compared with disposable e-cigarette users, e-liquid users were less likely to perceive smoking as cheaper (OR = 0.26, *p* < 0.01). Adult e-cigarette users who lived in a state with higher e-cigarette taxes (per ml) were more likely to perceive vaping as more expensive than smoking (OR = 1.43, *p* < 0.01). Neither cigarette use status nor state cigarette taxes were associated with adult vapers’ cost comparison between cigarettes and e-cigarettes.

**Table 2.** Associations between cost comparison and tobacco use and socio-demographics, ordered logit regression.

| Dependent variable: How do you compare the price of ECs to the price of CIGs? (1=less expensive, 2=same, 3=more expensive) | Odds ratio | 95% CI |
| --- | --- | --- |
| *EC use frequency (Reference group: Some days)* |  |  |
| Every day | 0.55* | [0.31,0.99] |
| *Cigarette use frequency (Reference group: Not at all)* |  |  |
| Some days | 1.36 | [0.76,2.43] |
| Every day | 2.21 | [0.89,5.47] |
| *Product type (Reference group: Disposable only)* |  |  |
| E-liquids only | 0.26** | [0.12,0.60] |
| Pre-filled pods only | 1.03 | [0.55,1.96] |
| Multiple | 0.79 | [0.45,1.41] |
| State EC tax | 1.43** | [1.10,1.87] |
| State CIG tax | 0.99 | [0.76,1.28] |
| *Sex (Reference group: Male)* |  |  |
| Female | 1.00 | [0.62,1.61] |
| *Age (Reference group: 18-29)* |  |  |
| 30-44 | 0.56 | [0.28,1.14] |
| 45-59 | 0.23** | [0.09,0.58] |
| 60+ | 0.32* | [0.11,0.88] |
| *Race/Ethnicity (Reference group: Non-Hispanic White)* |  |  |
| Non-Hispanic Black | 1.50 | [0.30,7.38] |
| Non-Hispanic other race | 0.95 | [0.30,3.06] |
| Hispanic | 1.37 | [0.65,2.88] |
| *Education attainment (Reference group: less than high school)* |  |  |
| High school | 0.66 | [0.27,1.58] |
| Some college or associate degree | 0.99 | [0.38,2.56] |
| Bachelor's Degree or more | 0.90 | [0.33,2.47] |
| *Employment status (Reference group: Working full-time)* |  |  |
| Working part-time | 1.38 | [0.55,3.48] |
| Not working | 1.31 | [0.63,2.73] |
| *Marital status (Reference group: Married)* |  |  |
| Not married | 1.46 | [0.90,2.38] |
| Number of adults in household | 1.24 | [0.96,1.59] |
| *Income (Reference group: Less than \$25,000)* | | |
| \$25,000 to \$49,999 | 0.49* | [0.24,1.00] |
| \$50,000 to \$74,999 | 0.92 | [0.45,1.87] |
| \$75,000 or more | 1.07 | [0.57,2.00] |
| *Housing Type (Reference group: One-family house)* |  |  |
| Condo/Townhouse attached | 1.77 | [0.64,4.89] |
| Apartment complex | 1.16 | [0.75,1.78] |
| Other | 0.84 | [0.36,1.97] |
| N | 720 |  |
| AIC | 1260.52 |  |
| BIC | 1402.47 |  |

EC: e-cigarette. CIG: cigarette. 95% CI: 95% confidence interval. \* $p < 0.05$ , \*\* $p < 0.01$ , \*\*\* $p < 0.001$ . Two cut points that indicated where the latent variable of this regression was cut to create the three ordered groups in our data (i.e., less expensive, same, more expensive) were estimated to be 2.57 [0.06,113.14] and 12.80 [0.27,616.62], respectively. The estimated odds ratio of the predicted probability of knowing the relative price were 4.00 [0.02,667.51]. The predicted probability was attained from the first stage in equation 1 of which the regression results are reported in Table A2.

Figures 1a and 1b show preferred budget or expenditure frequency (weekly vs. monthly) by tobacco use status (dual use or exclusive e-cigarette use) and use frequency (someday or everyday use). Dual users of cigarettes and e-cigarettes were 4% more likely to prefer reporting weekly budget on e-cigarettes compared with exclusive users of e-cigarettes, though this difference was not statistically significant. Everyday e-cigarette users were 16% more likely to prefer reporting weekly budget on e-cigarettes, relative to someday e-cigarette users (*p* < 0.01). Figure 2 presents frequency of cigarette budgeting and expenditures among dual users, and 61% of them preferred to report weekly cigarette budget. In addition, everyday smokers were 34% more likely to prefer reporting weekly cigarette budget (*p* < 0.001).

**Figure 1.**
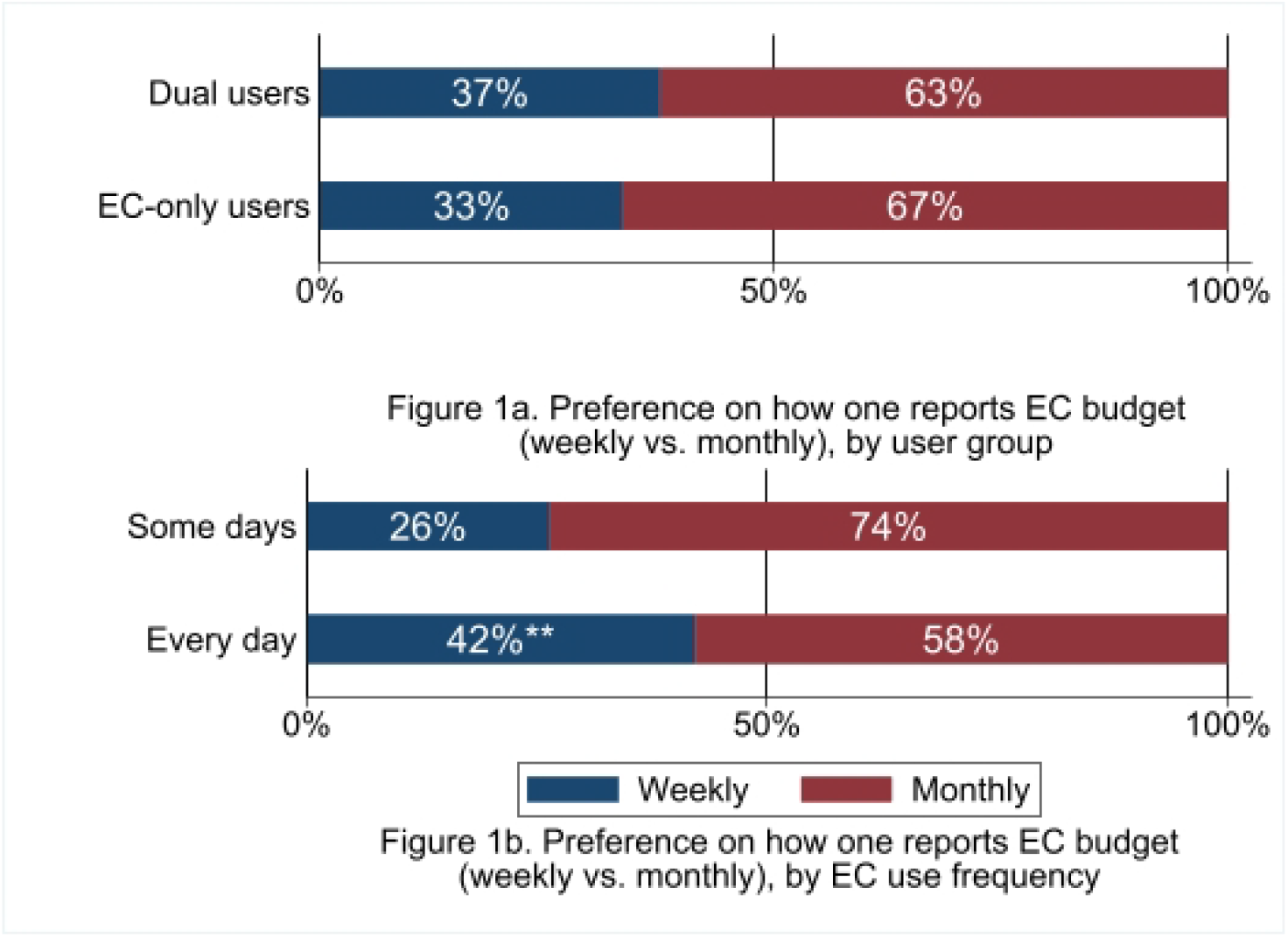
Preference on how one reports EC budget (weekly vs. monthly) (N=776)

**Figure 2.**
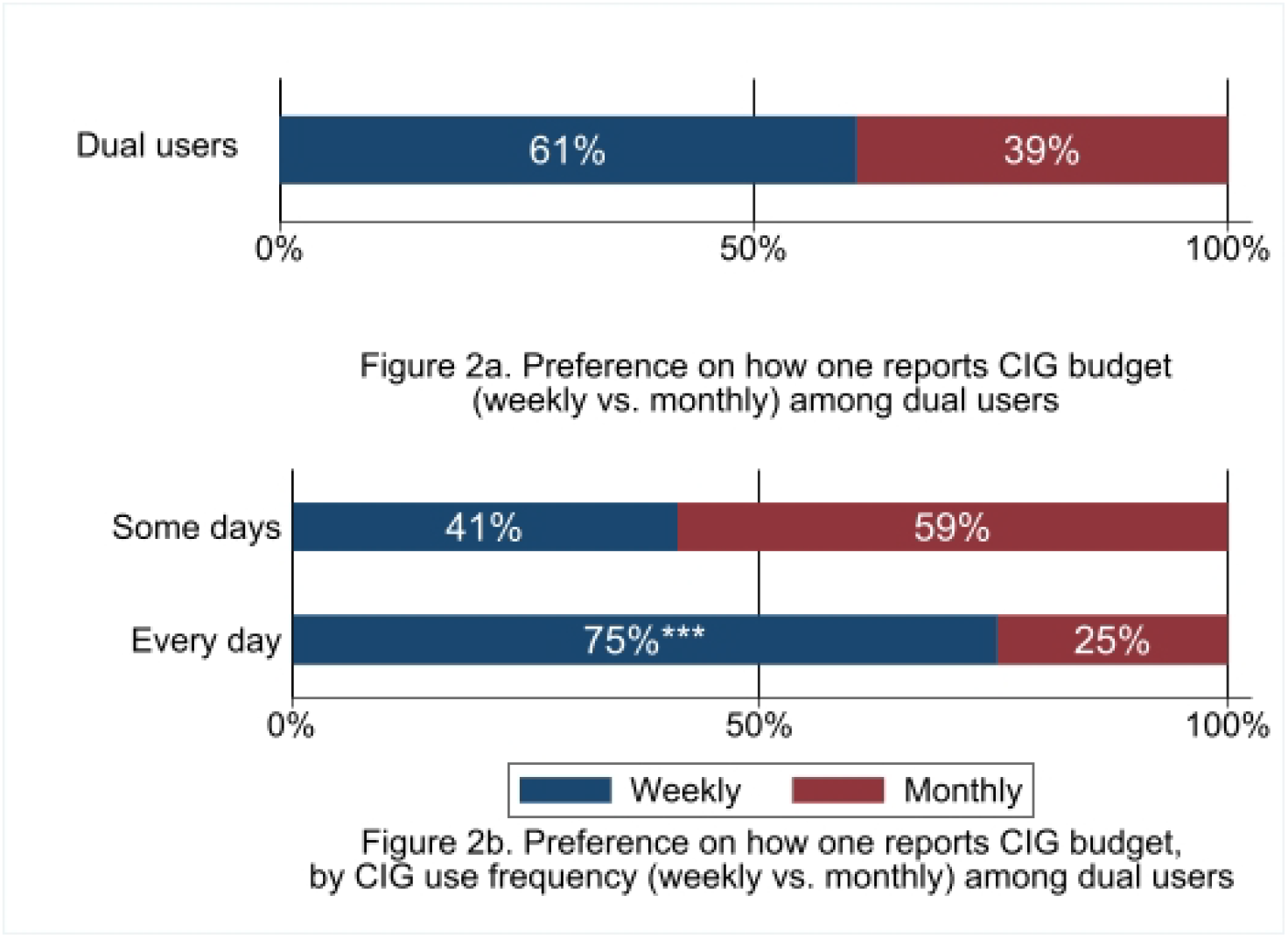
Preference on how one reports CIG budget (weekly vs. monthly) (N=268) EC: e-cigarette. CIG: cigarette. Dual users: Adult (18+) e-cigarette users who smoke cigarettes. **(***) in Figure 1b (Figure 2b) indicates that more regular users of EC(CIG) prefer budget weekly over monthly than less regular EC(CIG) users at 1% (0.1%) significance level. Alternatively, less regular EC(CIG) users prefer budget monthly over weekly than more regular EC(CIG) users since preference on weekly or monthly reporting is binary. In Figure 1a, dual users’ preferences of weekly over monthly were not statistically different from EC-only users. Each observation was weighted using the 2021 National Health Interview Survey as a benchmark, to ensure that the sample was representative of adult e-cigarette user population in the US.

Results from the expenditure analysis are shown in Table 3. Columns (1), (3) and (5) present findings without including cost comparison as an explanatory variable, whereas columns (2), (4) and (6) show findings when including cost comparison in the equations, to understand whether participants’ cost comparison between e-cigarettes and cigarettes mitigated the influences of other factors on their expenditures.

**Table 3.** Estimated coefficients from expenditure analysis using ordinary least square method.

|  | EC expenditure |  | CIG expenditure |  | Total expenditure |  |
| --- | --- | --- | --- | --- | --- | --- |
|  | (1) | (2) | (3) | (4) | (5) | (6) |
|  | No | Yes | No | Yes | No | Yes |
| *Cost comparison (Reference group: About the same)* |  |  |  |  |  |  |
| ECs are less expensive |  | -0.27* |  | 0.34 |  | -0.22* |
|  |  | (0.10) |  | (0.22) |  | (0.10) |
| ECs are more expensive |  | -0.10 |  | -0.18 |  | -0.16 |
|  |  | (0.14) |  | (0.18) |  | (0.13) |
| Don't know |  | -0.28** |  | -0.26 |  | -0.35** |
|  |  | (0.09) |  | (0.30) |  | (0.11) |
| *EC use frequency (Reference group: Some days)* |  |  |  |  |  |  |
| Every day | 0.69*** | 0.70*** | -0.21 | -0.29 | 0.53*** | 0.53*** |
|  | (0.08) | (0.08) | (0.16) | (0.16) | (0.09) | (0.09) |
| *CIG use frequency (Reference group is everyday use for columns 3-4)* |  |  |  |  |  |  |
| Some days | 0.22 | 0.19 | -1.12*** | -1.14*** | 0.75*** | 0.72*** |
|  | (0.13) | (0.14) | (0.14) | (0.13) | (0.12) | (0.12) |
| Every day | 0.16 | 0.11 | - | - | 1.46*** | 1.42*** |
|  | (0.09) | (0.09) |  |  | (0.11) | (0.11) |
| *Product type (Reference group: Disposable only)* |  |  |  |  |  |  |
| E-liquids only | -0.37** | -0.33** | 0.17 | 0.03 | -0.30* | -0.28* |
|  | (0.12) | (0.11) | (0.20) | (0.21) | (0.12) | (0.12) |
| Pre-filled pods only | 0.18 | 0.18 | -0.20 | -0.13 | 0.11 | 0.12 |
|  | (0.14) | (0.14) | (0.25) | (0.22) | (0.15) | (0.15) |
| Multiple | 0.08 | 0.09 | 0.25 | 0.25 | 0.08 | 0.10 |
|  | (0.12) | (0.11) | (0.22) | (0.21) | (0.12) | (0.11) |
| State EC tax | 0.09 | 0.08 | -0.13 | -0.10 | 0.08 | 0.08 |
|  | (0.05) | (0.05) | (0.08) | (0.08) | (0.06) | (0.06) |
| State CIG tax | 0.00 | 0.00 | 0.10 | 0.13 | 0.02 | 0.02 |
|  | (0.07) | (0.07) | (0.07) | (0.06) | (0.07) | (0.07) |
| *Sex (Reference group: Male)* |  |  |  |  |  |  |
| Female | -0.24** | -0.24* | 0.18 | 0.12 | -0.16 | -0.16 |
|  | (0.09) | (0.09) | (0.12) | (0.13) | (0.08) | (0.08) |
| *Age (Reference group: 18-29)* |  |  |  |  |  |  |
| 30-44 | 0.14 | 0.12 | 0.43* | 0.38 | 0.17 | 0.14 |
|  | (0.11) | (0.11) | (0.21) | (0.19) | (0.12) | (0.11) |
| 45-59 | -0.20 | -0.17 | 0.35 | 0.23 | -0.11 | -0.10 |
|  | (0.13) | (0.13) | (0.21) | (0.21) | (0.12) | (0.12) |
| 60+ | -0.33* | -0.31 | 0.53* | 0.42 | -0.19 | -0.20 |
|  | (0.16) | (0.17) | (0.24) | (0.24) | (0.18) | (0.19) |
| *Race/Ethnicity (Reference group: Non-Hispanic White)* |  |  |  |  |  |  |
| Non-Hispanic Black | 0.15 | 0.13 | -0.29 | -0.28 | 0.10 | 0.08 |
|  | (0.13) | (0.12) | (0.22) | (0.19) | (0.14) | (0.13) |
| Non-Hispanic other race | 0.09 | 0.12 | 0.35 | 0.34 | 0.13 | 0.16 |
|  | (0.18) | (0.18) | (0.30) | (0.26) | (0.17) | (0.16) |
| Hispanic | -0.11 | -0.16 | 0.24 | 0.23 | -0.11 | -0.14 |
|  | (0.19) | (0.19) | (0.16) | (0.14) | (0.18) | (0.18) |
| *Education attainment (Reference group: less than high school)* |  |  |  |  |  |  |
| High school | -0.12 | -0.12 | -0.07 | -0.12 | -0.06 | -0.07 |
|  | (0.11) | (0.11) | (0.20) | (0.19) | (0.11) | (0.12) |
| Some college or associate degree | -0.37** | -0.39** | -0.23 | -0.29 | -0.30* | -0.32* |
|  | (0.13) | (0.13) | (0.24) | (0.25) | (0.13) | (0.14) |
| Bachelor's Degree or more | -0.33* | -0.36* | -0.27 | -0.32 | -0.31* | -0.34* |
|  | (0.15) | (0.16) | (0.31) | (0.32) | (0.15) | (0.16) |
| *Employment Status (Reference group: Working full-time)* |  |  |  |  |  |  |
| Working part-time | -0.15 | -0.14 | -0.08 | -0.10 | -0.10 | -0.08 |
|  | (0.13) | (0.13) | (0.23) | (0.25) | (0.13) | (0.13) |
| Not working | -0.17 | -0.19 | -0.42* | -0.42* | -0.15 | -0.17 |
|  | (0.14) | (0.15) | (0.18) | (0.18) | (0.14) | (0.15) |
| *Marital Status (Reference group: Married)* |  |  |  |  |  |  |
| Not married | 0.01 | 0.02 | 0.07 | 0.08 | 0.04 | 0.06 |
|  | (0.09) | (0.08) | (0.15) | (0.14) | (0.10) | (0.09) |
| Number of adults in household | 0.05 | 0.03 | -0.06 | -0.07 | 0.03 | 0.02 |
|  | (0.04) | (0.04) | (0.06) | (0.07) | (0.04) | (0.04) |
| *Income (Reference group: Less than \$25,000)* | | | | | | |
| \$25,000 to \$49,999 | 0.07 | 0.09 | 0.04 | -0.03 | 0.03 | 0.03 |
|  | (0.13) | (0.13) | (0.24) | (0.23) | (0.13) | (0.13) |
| \$50,000 to \$74,999K | -0.05 | -0.06 | -0.14 | -0.17 | -0.04 | -0.05 |
|  | (0.16) | (0.16) | (0.35) | (0.36) | (0.16) | (0.17) |
| \$75,000 or more | -0.01 | -0.00 | 0.12 | 0.11 | 0.01 | 0.02 |
|  | (0.13) | (0.14) | (0.27) | (0.26) | (0.16) | (0.17) |
| *Housing type (Reference group: One-family house)* |  |  |  |  |  |  |
| Condo/Townhouse attached | -0.27* | -0.27* | -0.15 | -0.13 | -0.11 | -0.11 |
|  | (0.12) | (0.12) | (0.37) | (0.34) | (0.13) | (0.13) |
| Apartment complex | -0.17 | -0.20* | -0.21 | -0.22 | -0.16 | -0.20 |
|  | (0.10) | (0.10) | (0.19) | (0.22) | (0.10) | (0.10) |
| Other | -0.22 | -0.27 | -0.28 | -0.33 | -0.19 | -0.26 |
|  | (0.14) | (0.14) | (0.23) | (0.22) | (0.16) | (0.15) |
| N | 776 | 776 | 268 | 268 | 776 | 776 |
| AIC | 1975.76 | 1968.91 | 732.43 | 724.12 | 1946.17 | 1939.17 |
| BIC | 2110.73 | 2117.84 | 832.98 | 835.44 | 2081.14 | 2088.10 |

EC: e-cigarette. CIG: cigarette. \* $p < 0.05$ , \*\* $p < 0.01$ , \*\*\* $p < 0.001$ . Standard errors clustered at the state level are in parentheses. Expenditures are monthly or converted monthly by multiplying weekly expenditures by 4 and taken natural logs. Zero-value EC (CIG) expenditures were excluded from EC (CIG) expenditure analyses. Total expenditure is the sum of EC and CIG expenditures, excluding individuals with zero expenditure on ECs.

Regardless of controlling for cost comparison or not, everyday e-cigarette users spent 69-70% more on e-cigarettes (*p* < 0.001) and 53% more on e-cigarettes and cigarettes combined if they were dual users (*p* < 0.001), compared with someday e-cigarette users. E-liquid users spent 33-37% less on e-cigarettes and 28-30% less on e-cigarettes and cigarettes combined if they were dual users, relative to disposable e-cigarette users. Compared with adult vapers who had less than high school education, those with some college education, associate degree or bachelor’s degree spent 33-39% less on e-cigarettes and 30-34% less on cigarettes and e-cigarettes combined if they were dual users. Compared to adults who lived in a one-family house, those who lived in a condo, townhouse or apartment complex spent less on e-cigarettes. In addition, female participants spent 24% less on e-cigarettes than male participants.

Among dual users of cigarettes and e-cigarettes, someday smokers spent less on cigarettes than everyday smokes. Total expenditures (cigarettes and e-cigarettes combined) of dual users who were someday smokers or everyday smokers were 72-75% and 1.4 times more than those of exclusive e-cigarette users, respectively. Compared to dual users aged 18-29, those who were 30-44 or 60+ spent more on cigarettes. Among dual users, those who were not working spent 42% less on cigarettes than full-time workers.

Finally, state-level cigarette and e-cigarette taxes were not associated with e-cigarette or cigarette expenditures, regardless of controlling for cost comparison. The associations, AIC and BIC were very similar across models with or without cost comparison as a control variable. This finding suggests that cost comparison did not mediate any potential associations between expenditures and explanatory variables in the regressions. Nonetheless, cost comparisons were significantly associated with e-cigarette expenditures alone, and total expenditures (of cigarettes and e-cigarettes combined), suggesting their possible behavioral impacts.

## DISCUSSION

Our study adds to the tobacco control literature by presenting crucial evidence on e-cigarette and cigarette cost comparison and expenditures using data from a nationally representative sample of adult e-cigarette users aged 18+ in the U.S. We found that monthly e-cigarette spending was $82.22 and cigarette spending was $118.77, with the combined cigarette and e-cigarette spending amounting to 217.11 dollars for dual users who purchased both. Considering that average household income was less than $6,250 per month (i.e., < $75,000 per year) in our sample, this finding implies that over 1.3% income was spent on e-cigarettes alone, and for dual users, 2% was spent on cigarettes. These percentages could be significantly higher for users with low income (e.g., less than $40,000).

Consistent with the growing popularity of disposables since flavors were banned in open systems,(Do et al., 2023; Hammond et al., 2022) disposables were also the most popular e-cigarette product type among people who exclusively used one e-cigarette model or type (45%), followed by exclusive users of e-liquids (37%) and refill cartridges/pods (18%). In addition, 26% of adult vapers reported using multiple e-cigarette models or types. Existing literature suggests that while young adult vapers prefer pods and disposables, older adults may prefer tank or mod systems.(Tillery et al., 2023) In addition, different device types may have different appeals, and their potentials as cigarette substitute as well as health risks might also differ.(Hajek et al., 2018) Our study shows that a majority of vapers exclusively used one e-cigarette model or type, suggesting the importance of evaluating e-cigarette use and health consequences by device types.

Interestingly, we found that adult vapers who lived in states with higher e-cigarette excise taxes were more likely to perceive vaping as more costly than smoking, which is consistent with existing literature that shows higher e-cigarette taxes reduce e-cigarette use and sales.(Cotti et al., 2022; Reiter et al., 2024; Yan et al., 2023) In contrast, cigarette taxes were not significantly associated with cost comparison among adult vapers. Our findings are different from a previous study that shows cigarettes taxes are associated with perceiving cigarettes as more costly than e-cigarettes among US adult smokers and e-cigarette users.(He et al., 2024) These differences may be explained by sampling differences between the two studies since the prior study used a national sample primarily consisting of adult smokers (78%)(He et al., 2024) whereas our sample exclusively consists of vapers (70% exclusive e-cigarette users and 30% dual users). It is not surprising that cigarette taxes influence cost comparison among smokers and e-cigarette taxes influence cost comparison among vapers, but not vice versa.

Despite the differences in the associations between taxes and cost comparison, our study and prior research converge in showing that vapers who used tank systems with e-liquids perceived lower costs of vaping compared to other device types. This may be desired because there is research suggesting that vapers who successfully quit smoking prefer tank systems with e-liquids to other types.(Chen et al., 2016; Dawkins et al., 2013) Nonetheless, given the different tax burdens on difference device types,(Shang et al., 2023) more research is needed to understand the roles of economic costs across device types in shaping vaping and smoking behaviors.

Furthermore, our cost comparison analysis illustrated vapers aged 45+ perceived lower costs of e-cigarettes than younger adults, and that vapers with 25K to less than 50K household income perceived lower costs of e-cigarettes compared to people with a household income less than 25K. These findings are in general in line with existing knowledge that younger people and tobacco users of lower SES are more sensitive to prices.(Cruces et al., 2022; Golden et al., 2020)

Our expenditure analysis further confirmed the findings in cost comparison analysis: compared to those who perceived cigarettes and e-cigarettes as costing the same, vapers who perceived lower e-cigarette costs also spent less on e-cigarettes, and if they were dual users, spent less on e-cigarettes and cigarettes combined, after controlling for use frequencies. Similarly, e-liquid users spent less than users of other e-cigarette product types. These findings suggest that cost comparison and expenditures are related concepts to consumers, measuring several factors including actual costs (e.g., taxes), use patterns, and SES. From this perspective, both cost comparison and expenditure measures are constructs more similar to affordability than to actual costs or pricing. Future research is needed to elucidate their relationships and examine how use behaviors are jointly determined by device types, perceived costs, and budget/expenditure constraints.

The expenditure analysis further showed that female e-cigarette users, those who were at least 60 years, and people with at least some college education spent significantly less than their counterparts on e-cigarettes, after adjusting for frequency of use, income, cost comparison, etc. These findings corroborate with the relatively low prevalence of use among these subpopulations reported by surveillance data. Interestingly, vapers who lived in a condo or townhouse spent less than those who lived in a one-family house, after controlling for household income. This finding suggests possible effects of tobacco-free policies (commonly imposed in shared housing) on reducing e-cigarette spending.(Azagba et al., 2020)

Finally, we found that budgeting window and spending frequency (weekly vs. monthly) were significantly associated with use frequency: everyday users preferred to report weekly budget whereas someday users preferred to report monthly budget. Future tobacco surveys may consider this difference when eliciting spending-related questions. Nonetheless, the percentage of vapers reporting weekly purchases of e-cigarettes (42%) was much lower than that of dual users reporting weekly purchases of cigarettes (75%), showing considerable difference in purchasing frequencies between cigarettes and e-cigarettes.

Our study is not without limitations. First, the conclusion is based on cross-sectional data from a single survey. Future research using repeated cross-sectional or longitudinal data are needed to ascertain the relationships. Second, cost comparison and expenditures are based on self-reported information and may contain recall or measurement errors. Third, although we collected expenditure data and had detailed information on use frequencies and household income, we have not used Deaton or household expenditure model to estimate own and cross price elasticities of demand for cigarettes and e-cigarettes, which we plan to assess in future studies.(Jackson et al., 2019; John et al., 2023)

## CONCLUSIONS

Adult e-cigarette users in the U.S. on average spends $82 on e-cigarettes and dual users spend additional $119 on cigarettes per month. A majority of adult e-cigarette users exclusively used one device type, and among them, disposables were the most popular. State excise taxes on e-cigarettes, device type, and age were associated with adult users’ cost perception of vaping relative to smoking. Compared to adult users who perceived vaping and smoking as costing the same, those who perceived vaping as less costly (than smoking) also spent less on vaping. Age, sex, education attainment, housing type and device type were associated with adult users’ spending on vaping. Daily e-cigarette users tended to budget expenditures on a weekly basis whereas someday users tended to budget expenditures on a monthly basis. Future research is warranted to understand how e-cigarette use behaviors among US adults are jointly determined by device types, perceived costs, and budget/expenditure constraints.

## Data Availability

All data produced in the present study are available upon reasonable request to the authors

## Ethics Statement

This study was approved by The Ohio State University Institutional Review Board (Study ID: 2021E0460).

## Acknowledgment

We thank Yousef Alish for his assistance with this research project.

## Author contribution statement

SM – validation, investigation, writing - original draft, writing - review & editing, visualization; QY - investigation, project administration, writing - review & editing; SA – software, formal analysis, data curation, writing - original draft, visualization; HP - investigation, writing - review & editing; YH – investigation, writing - review & editing; JB – conceptualization, methodology, writing - review & editing, supervision; CS – conceptualization, methodology, software, validation, investigation, resources, writing - review & editing, supervision, funding acquisition

