## Appendix for "Cost Comparison and Spending on Tobacco Products: Evidence from A Nationally Representative Sample of Adult E-Cigarette Users"

**Table A1.** Summary statistics of adult EC users' demographic characteristics

| *Variable* | Cost comparison analysis | | Expenditure analysis | |
|---|---|---|---|---|
| | Frequency | Weighted percentage /mean | Frequency | Weighted percentage /mean |
| *Number of adults (18+) in the household* | 801 | 2.31 | 776 | 2.32 |
| *Age* | | | | |
| 18-29 | 131 | 42.49% | 125 | 41.37% |
| 30-44 | 287 | 32.53% | 279 | 33.13% |
| 45-59 | 225 | 15.89% | 218 | 16.08% |
| 60+ | 158 | 9.09% | 154 | 9.42% |
| *Sex* | | | | |
| Male | 328 | 53.79% | 316 | 53.71% |
| Female | 473 | 46.21% | 460 | 46.29% |
| *Marital status* | | | | |
| Married | 321 | 31.86% | 316 | 33.21% |
| Not married (divorced, separated, or single) | 480 | 68.14% | 460 | 66.79% |
| *Race/Ethnicity* | | | | |
| Non-Hispanic White | 611 | 73.21% | 591 | 73.64% |
| Non-Hispanic Black | 49 | 6.31% | 46 | 6.05% |
| Other/Two or more races | 54 | 7.86% | 54 | 8.26% |
| Hispanic | 87 | 12.62% | 85 | 12.05% |
| *Education* | | | | |
| Less than high school | 46 | 7.21% | 41 | 7.16% |
| High school graduate or equivalent | 167 | 36.00% | 161 | 36.00% |
| Some college or associate degree | 356 | 35.83% | 350 | 35.34% |
| Bachelor's Degree or higher | 232 | 20.96% | 224 | 21.50% |
| *Current employment status* | | | | |
| Working full-time | 418 | 54.33% | 409 | 55.76% |
| Working part-time | 134 | 19.11% | 126 | 17.35% |
| Not working | 249 | 26.56% | 241 | 26.88% |
| *Household income (before tax) in the past 12 months* | | | | |
| Less than $25,000 | 177 | 16.81% | 166 | 14.98% |
| $25,000 to $49,999 | 180 | 20.40% | 175 | 20.21% |
| $50,000 to $74,999 | 149 | 18.70% | 142 | 19.12% |
| $75,000 or more | 295 | 44.09% | 293 | 45.70% |
| *Housing type* | | | | |
| One-family house | 507 | 64.83% | 497 | 66.95% |
| One-family condo/townhouse | 75 | 9.23% | 72 | 7.56% |
| Apartments | 160 | 18.94% | 151 | 19.21% |
| Other (mobile home, boat, RV, van) | 59 | 7.00% | 56 | 6.28% |
| *N* | 801 | | 776 | |

The average number of adults in the household (continuous variable) and the percentage of each category (in categorical variables) are all weighted using individual weights. The individual weights were designed so that our sample was representative of adult (18+) e-cigarette user population in the US. The benchmark survey used in the weighting process was the 2021 National Health Interview Survey. The total number of study participants were 808, and 7 who did not reveal their e-cigarette product type of use or refused to answer the relative prices of e-cigarettes and cigarettes were excluded, resulting in a total of 801 participants used in the cost comparison analysis. For the expenditure analysis, 25 participants who reported their spending on e-cigarettes as zero were further excluded, resulting in 776. Please see the "Recruitment" subsection within the "Method" section for more details about weighting and participant exclusion criterion.

**Table A2**. The first stage of cost comparison analysis using logit regression

| Dependent variable: Do you know the relative cost of e-cigarettes and cigarettes? (0=Don't know, 1=Know) | Odds ratio | 95% CI |
|---|---|---|
| *EC use frequency (Reference group: Some days)* | | |
| Every day | 1.70 | [0.82,3.53] |
| *CIG use frequency (Reference group: Not at all)* | | |
| Some days | 1.62 | [0.62,4.18] |
| Every day | 3.36 | [0.97,11.61] |
| *Product type (Reference group: Disposable only)* | | |
| E-liquids only | 2.24 | [0.79,6.31] |
| Pre-filled pods only | 0.51 | [0.20,1.33] |
| Multiple | 0.94 | [0.41,2.15] |
| State e-cigarette tax | 0.93 | [0.52,1.64] |
| State cigarette tax | 1.15 | [0.78,1.69] |
| *Sex (Reference group: Male)* | | |
| Female | 1.13 | [0.59,2.19] |
| *Age (Reference group: 18-29)* | | |
| 30-44 | 1.79 | [0.72,4.42] |
| 45-59 | 2.07 | [0.80,5.38] |
| 60+ | 4.00* | [1.23,12.98] |
| *Race/Ethnicity (Reference group: Non-Hispanic White)* | | |
| Non-Hispanic Black | 2.73 | [0.48,15.48] |
| Non-Hispanic other race | 0.35 | [0.12,1.07] |
| Hispanic | 1.91 | [0.56,6.45] |
| *Education attainment (Reference group: less than high school)* | | |
| High school | 1.15 | [0.26,4.99] |
| Some college or associate degree | 2.65 | [0.61,11.60] |
| Bachelor's Degree or more | 2.20 | [0.45,10.77] |
| *Employment Status (Reference group: Working full-time)* | | |
| Working part-time | 0.61 | [0.23,1.64] |
| Not working | 0.93 | [0.38,2.29] |
| *Marital Status (Reference group: Married)* | | |
| Not married | 0.46 | [0.18,1.14] |
| Number of adults in household | 1.25 | [0.85,1.82] |
| *Income (Reference group: Less than $25,000)* | | |
| $25,000 to $49,999 | 1.86 | [0.63,5.52] |
| $50,000 to $74,999 | 1.95 | [0.65,5.81] |
| $75,000 or more | 0.85 | [0.29,2.48] |
| *Housing Type (Reference group: One-family house)* | | |
| Condo/Townhouse attached | 1.19 | [0.33,4.29] |
| Apartment complex | 1.06 | [0.43,2.62] |
| Other | 4.22 | [0.92,19.42] |
| N | 801 | |

EC: e-cigarette. CIG: cigarette. 95% CI: 95% confidence interval. AIC: 669.87. BIC: 805.76. * $p < 0.05$, ** $p < 0.01$, *** $p < 0.001$.
